# A Rapid review on the COVID-19 Pandemic’s Global Impact on Breast Cancer Screening Participation Rates and Volumes from January-December 2020

**DOI:** 10.1101/2023.02.06.23285513

**Authors:** Reagan Lee, Wei Xu, Marshall Dozier, Ruth McQuillan, Evropi Theodoratou, Jonine D. Figueroa, UNCOVER and the International Partnership for Resilience in Cancer Systems (I-PaRCS), Breast Cancer Working Group 2

## Abstract

**Background:** COVID-19 has strained population breast mammography screening programs that aim to diagnose and treat breast cancers earlier. As the pandemic has affected countries differently, we aimed to quantify changes in breast screening volume and uptake during the first year of the COVID-19 pandemic.

**Methods:** We systematically searched Medline, the WHO (World Health Organization) COVID-19 database, and governmental databases. Studies covering January 2020 to March 2022 were included. We extracted and analyzed data regarding study methodology, screening volume and uptake. To assess for risk-of-bias, we used the Joanna Briggs Institute Critical Appraisal tool.

**Results:** Twenty-six cross-sectional descriptive studies were included out of 935 independent records. Reductions in screening volume and uptake rates were observed among eight countries. Changes in screening participation volume in five countries with national population-based screening ranged from -13% to –31%. Among two countries with limited population-based programs the decline ranged from -61% to -41%. Within the USA, population participation volumes varied ranging from +18% to -39% with suggestion of differences by insurance status (HMO, Medicare, and low-income programs). Almost all studies had high risk-of-bias due to insufficient statistical analysis and confounding factors.

**Discussion and Conclusion:** Extent of COVID-19-induced reduction in breast screening participation volume differed by region and data suggested potential differences by healthcare setting (e.g., national health insurance vs private health care). Recovery efforts should monitor access to screening and early diagnosis to determine if prevention services need strengthening to increase coverage of marginalized groups and reduce disparities.

## Introduction

Breast cancer is the most common cancer worldwide with 2.3 million women diagnosed and 685000 deaths in 2020 (WHO, 2022). Mammography-based screening programs allow for early detection of breast cancers, allowing for earlier intervention and disease stage that improves patient outcomes (IARC, 2022). Early detection and diagnosis from screening may reduce mortality up to 65% among breast cancer patients (Berry *et al*. 2005). Populations with a good uptake rate in screening programs can achieve a 90% 5-year survival rate in patients who received an early diagnosis attributed to screening (WHO, 2020).

The ongoing COVID-19 affected global health systems and has strained population breast mammography screening programs. Previous work on modelled evaluations and a focus on tumor staging and mortality as outcomes, suggested scenarios are likely to differ by region and organization of delivery of breast cancer screening (*Figueroa et al. Prev Med 2021*). In different countries, screening models vary from population-based to opportunistic screening (offered to patients in healthcare settings – more common in private healthcare) (IARC, 2016).

Here we aimed to quantify systematically breast screening participation rates before and after the COVID-19 including suspensions in nations with/without opportunistic screening programs. This was performed by investigating two primary study outcomes: changes in screening volume and participation rates.

## Methods

We performed a rapid review (Tricco et al. 2015), where systematic review processes were modified to facilitate project completion within a shortened timeframe. Searches were limited to two databases and English-language governmental grey literature.

### Literature search

RL ran a systematic search in on “Ovid MEDLINE(R) and In-Process, In-Data-Review & Other Non-Indexed Citations” Database and WHO COVID-19 Literature Database with entry date limits from 1 January 2020 to 12 March 2022. In brief, we performed the search with MeSH subject headers and free text terms for “COVID-19”, “Breast Neoplasms” and “Mass screening”. Our search strategies are listed in **Table 1**. We searched grey literature from government health websites known to have data from population-based screening programs. These consisted of the National Cancer Institute (USA), CDC (USA), NHS (National Healthcare Service) UK database, BreastScreen Australia and BreastScreen Aotearoa New Zealand. We further screened reference lists of the retrieved eligible publications to identify additional relevant studies. An English language restriction was placed on the searches. Deduplication was carried out as part of upload to Covidence systematic review software, Veritas Health Innovation, Melbourne, Australia. Available at www.covidence.org.

**Table 1:** Search strategies for rapid review of breast cancer participation and volume during Covid.

| Search String for Ovid MEDLINE(R) and In-Process, In-Data-Review & Other Non-Indexed |  |  |
| --- | --- | --- |
| Citations |  |  |
| Search Number | Search Domain | Search string in: [mp=title, book title, abstract, original title, name of substance word, subject heading word, floating sub-heading word, keyword heading word, organism supplementary concept word, protocol supplementary concept word, rare disease supplementary concept word, unique identifier, synonyms] |
| 1 | COVID-19 | (COVID-19 OR 2019 novel coronavirus disease OR 2019 novel coronavirus infection OR 2019 ncov disease OR 2019 ncov infection OR 2019-ncov disease OR 2019-ncov diseases OR 2019-ncov infection OR 2019-ncov infections OR covid 19 OR covid 19 pandemic OR covid 19 virus disease OR covid 19 virus infection OR covid-19 OR covid-19 pandemic OR covid-19 pandemics OR covid-19 virus disease OR covid-19 virus diseases OR covid-19 virus infection OR covid-19 virus infections OR covid19 OR coronavirus disease 19 OR coronavirus disease 2019 OR coronavirus disease-19 OR disease 2019, coronavirus OR sars cov 2 infection OR sars coronavirus 2 infection OR sars-cov-2 infection OR sars-cov-2 infections OR severe acute respiratory syndrome coronavirus 2 infection OR disease, 2019-ncov OR disease, covid-19 virus OR infection, 2019-ncov OR infection, covid-19 virus OR infection, sars-cov-2 OR pandemic, covid-19 OR virus disease, covid-19 OR virus infection, covid-19 OR Coronavirus, 2019 Novel OR ncov OR covid* OR coronavirus* OR SARS* OR severe acute respiratory syndrome OR coronavirus pandemic OR coronavirus disease pandemic) |
| 2 | Breast Cancer | (Breast Neoplasms OR Breast Carcinoma In Situ OR Carcinoma, Ductal, Breast OR Carcinoma, Lobular OR breast cancer OR breast carcinoma* OR breast malignant neoplasm* OR breast malignant tumor* OR breast neoplasm* OR breast tumor* OR cancer of breast? OR cancer of the breast? OR mammary carcinoma* OR mammary neoplasm* OR malignant neoplasm? of breast OR malignant tumor? of breast OR mammary cancer* OR neoplasm?, breast OR tumor?, breast OR cancer?, breast OR cancer?, mammary OR carcinoma?, human mammary OR carcinoma?, breast OR neoplasm?, human mammary OR breast carcinoma in situ OR lobular carcinoma in situ OR lcis, lobular carcinoma in situ OR mammary ductal carcinoma? OR carcinoma, ductal, breast OR carcinoma, infiltrating duct OR carcinoma, invasive ductal, breast OR carcinoma, mammary ductal OR carcinomas, infiltrating duct OR carcinomas, mammary ductal OR invasive ductal carcinoma, breast OR lobular carcinoma? OR carcinoma?, lobular OR breast* OR breast tumor? OR breast tumor? OR breast malignant tumor? OR breast malignan* OR mammary malignan* OR malignant tumor? of breast OR neoplasm? of breast OR breast neoplasm OR lcis) |
| 3 | Mass Screening | (Mass Screening OR Mass Chest X-ray OR Early Diagnosis OR Early Detection of Cancer OR Mammography OR screening* OR Ultrasonography, Mammary OR Ultrasonography OR mass chest x ray OR mass chest x-ray* OR mass chest xray* OR x-ray, mass chest OR x-rays, mass chest OR xray, mass chest OR xrays, mass chest OR disease early detection OR early detection of disease OR early diagnosis OR diagnosis, early OR cancer early detection OR cancer early diagnosis OR early detection of cancer OR early diagnosis of cancer OR digital breast tomosyntheses OR digital breast tomosynthesis OR x ray breast tomosynthesis OR x-ray breast tomosyntheses OR x-ray breast tomosynthesis OR breast tomosyntheses, digital OR breast tomosyntheses, x-ray OR breast tomosynthesis, digital OR breast tomosynthesis, x-ray OR breast tissue imaging OR mastography OR mass breast xray OR mass breast x-ray OR chest xray OR chest x-ray OR mammogra* OR program* OR ultrasonic* OR echograph* OR echotomograph* OR sonography* OR ultrasonograph* OR ultrasound* OR exam*) |
| 4 | Search String | 1 AND 2 AND 3 |
| 5 | Final Search String | limit 4 to english language |
| Search String for WHO COVID-19 Literature Database (updated to 12 March 2022) |  |  |
| Search Number | Search Concept | Title, Abstract, Subject |
| #1 | Breast Cancer | ((Breast Neoplasms) OR (Breast Carcinoma In Situ) OR (Carcinoma, Ductal, Breast) OR (Carcinoma, Lobular) OR (breast cancer*) OR (breast carcinoma*) OR (breast malignant neoplasm*) OR (breast malignant tumor*) OR (breast neoplasm*) OR (breast tumor*) OR (cancer of breast?) OR (cancer of the breast?) OR (mammary carcinoma*) OR (mammary neoplasm*) OR (malignant neoplasm? of breast) OR (malignant tumor? of breast) OR (mammary cancer*) OR (breast carcinoma in situ) OR (lobular carcinoma in situ) OR (mammary ductal carcinoma*) OR (breast ductal carcinoma*) OR (infiltrating duct carcinoma*) OR (invasive ductal carcinoma) OR (mammary ductal carcinoma*) OR (invasive ductal breast carcinoma) OR (lobular carcinoma*) OR (breast tumor*) OR (breast malignant tumor*) OR (breast malignan*) OR (mammary malignan*) OR (malignant tumor? of breast) OR (neoplasm? of breast) OR (lcis*)) |
| #2 | Screening | ((Mass Screening) OR (Mass Chest X-ray) OR (Early Diagnosis) OR (Early Detection of Cancer) OR (Mammography) OR (Ultrasonography, Mammary) OR (Ultrasonography) OR (national screening) OR (screening*) OR (mass chest x ray) OR (mass chest x-ray*) OR (mass chest xray*) OR (mass chest x-ray*) OR (disease early detection) OR (early detection of disease) OR (early diagnosis) OR (diagnosis, early) OR (cancer early detection) OR (cancer early diagnosis) OR (early detection of cancer) OR (early diagnosis of cancer) OR (digital breast tomosyntheses) OR (digital breast tomosynthesis) OR (x ray breast tomosynthesis) OR (x-ray breast tomosyntheses) OR (breast tomosynthesis*) OR (breast tissue imaging) OR (mastography) OR (mass breast xray) OR (mass breast x-ray) OR (chest xray) OR (chest x-ray) OR (mammogra*) OR (program*) OR (ultrasonic*) OR (echograph*) OR (ultrasonographic*) OR (sonography*) OR (echotomograph*) OR (ultrasound*) OR (exam*)) |
| #3 | Final Search string | #1 AND #2<br>English language filter |

### Inclusion and Exclusion Criteria

The Population, Interventions, Comparator, Outcomes, and Study Characteristics (PICOS) model (Schardt et al. 2007) was used to determine eligibility criteria. A pilot literature screen (n = 10) was performed by (RL with guidance from MD and JF) to confirm validity of criteria.

The population of focus are women eligible for breast cancer screening programs (population-based or opportunistic) globally or breast screening programs that are a part of the International Screening Cancer Network (ISCN). The intervention investigated involves the introduction of COVID-19 infection control measures.

These were assumed to be present globally due to worldwide prevalence of COVID-19 by March 2020 (chosen due to WHO’s declaration of a pandemic. The comparator involved comparing breast cancer screening statistics after COVID-19 related screening shutdown versus an analogous period in the previous years (e.g., comparing statistics in Australia from May-Sep 2020 against data from May-Sep 2018/2019) or any relevant period.

Outcomes assessed were volume of breast screening participation (“volume”) defined as total number of breast screening procedures; participation uptake rate (“uptake rate”) of breast screening program defined as the percentage of the eligible population who attend screening; and incidence of breast cancer diagnosis. These were compared between each comparator period.

Full-text, English-language primary papers or governmental published grey literature were included.

Studies with data entirely pertaining to diagnostic imaging were excluded or with future modelled data were excluded. All studies focused on women

Studies were required to have data on breast screening following the resumption of breast screening in countries with a screening shutdown.

### Title, Abstract, Full text Screen

Two reviewers (RL, JF) parallelly independently reviewed titles, abstracts, and subsequently full texts based on pre-defined inclusion and exclusion criteria. Deduplication of articles and screening was performed on Covidence. Conflict resolution was performed by discussion.

## Data Extraction

Data extraction for each article was conducted by a single reviewer (RL). A second reviewer (WX) then checked for eligibility of extracted data in 70% of the texts. Any conflicts were resolved by a third reviewer (JF). Data relevant to the evidence for population-based or opportunistic breast cancer screening programs during COVID-19 were extracted including citation details, publication type, study design, country, region, population, study setting, screening sample size, screening timeframe, screening volumes change (before/after COVID-19 infection control guidelines), screening participation rates change (before/after COVID-19 infection control guidelines), breast cancer incidence rates. A standardized data extraction form was created and piloted for extraction of primary outcome measures.

### Risk of Bias Assessment

All studies included had cross-sectional designs. We used the Joanna Briggs Institute Critical appraisal tool for cross-sectional studies to assess the risk of bias of each article (Joanna Briggs Institute, 2022). The JBI checklist is available in Table 4. Risk-of-bias for each article was assessed by a single reviewer [RL], and a second reviewer [WX] cross-assessed the results and verified all related judgement and rationales. Discrepancies were resolved through discussion and a joint reassessment of studies.

**Table 2.**
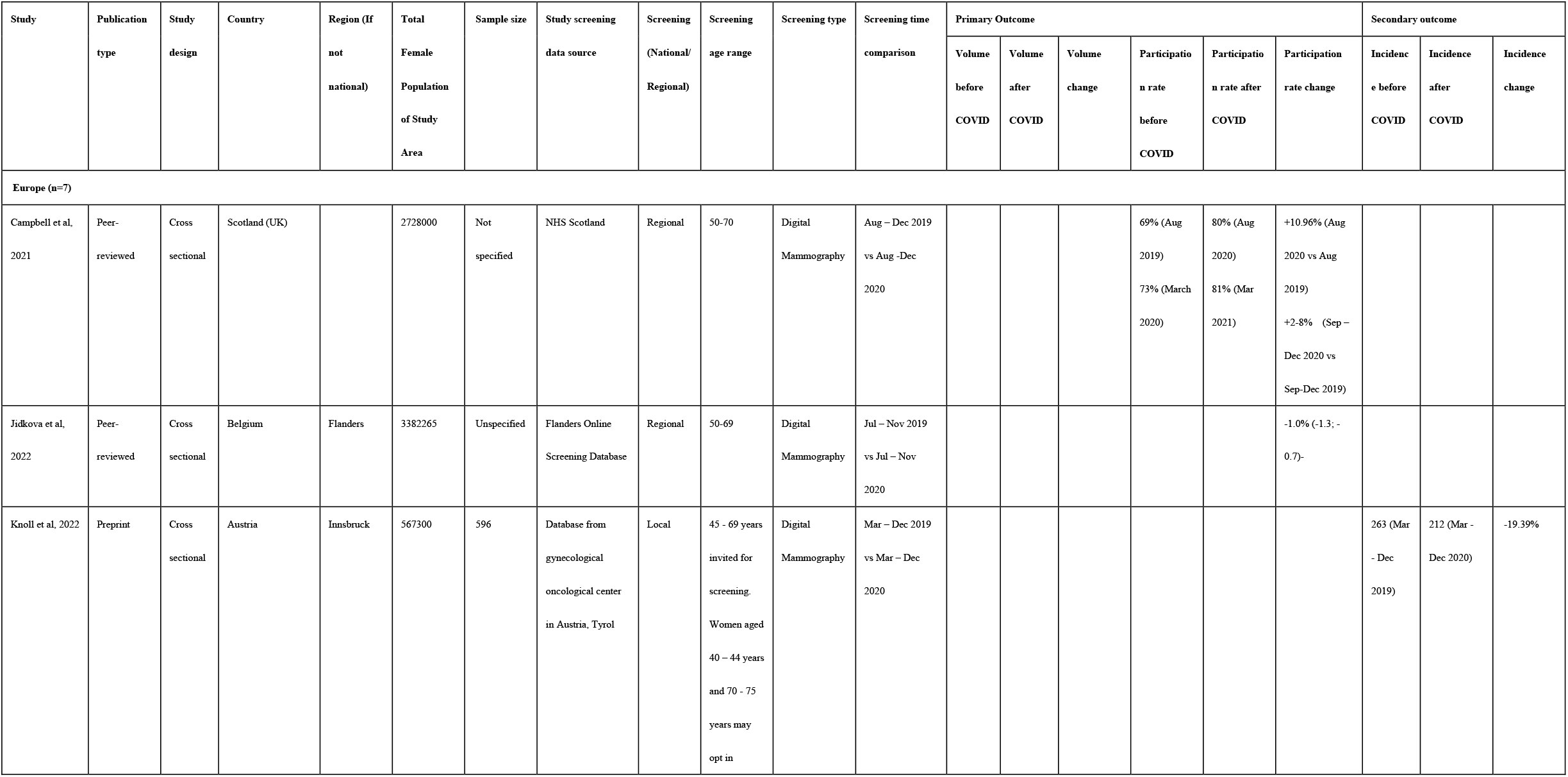
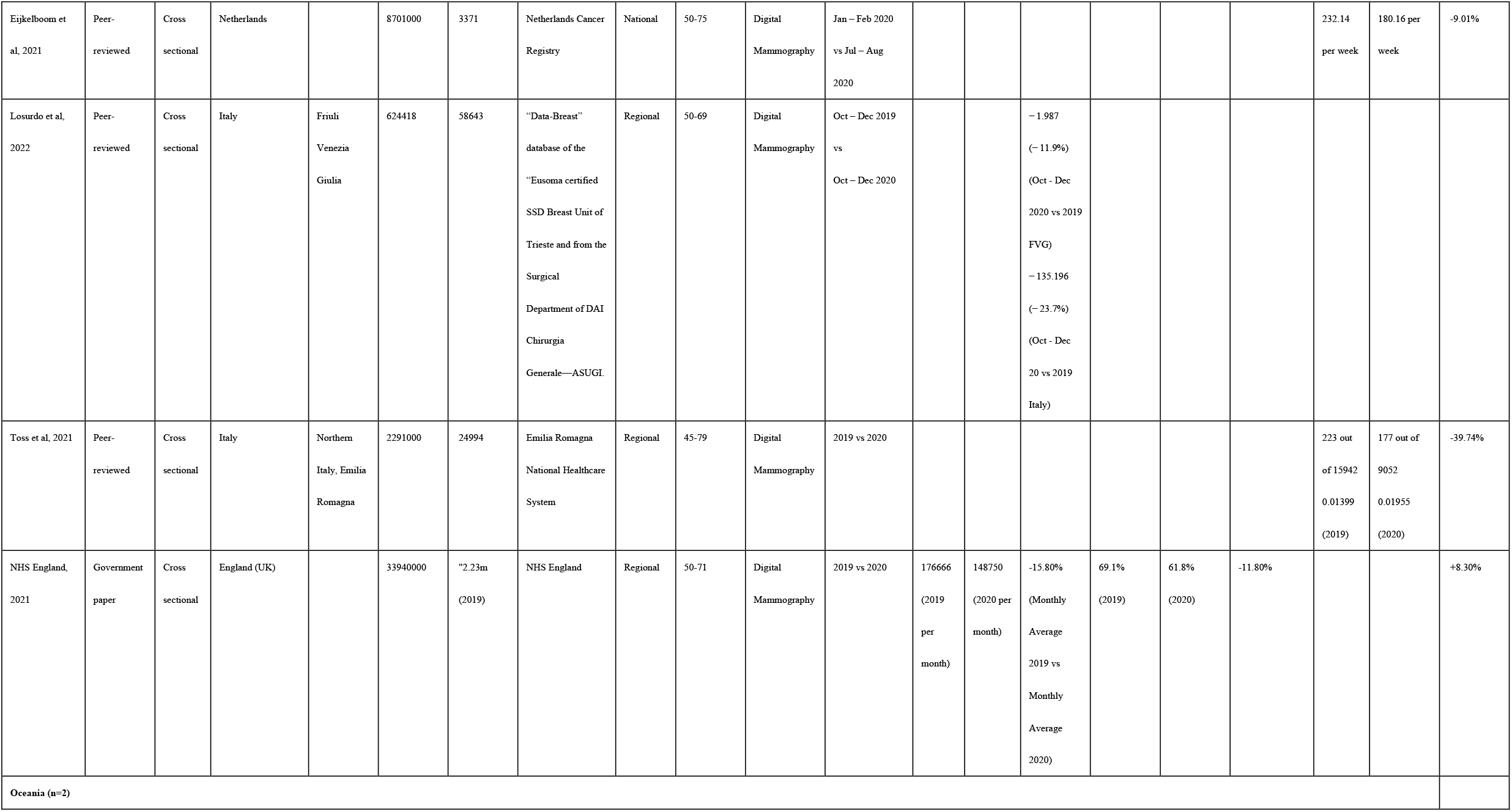
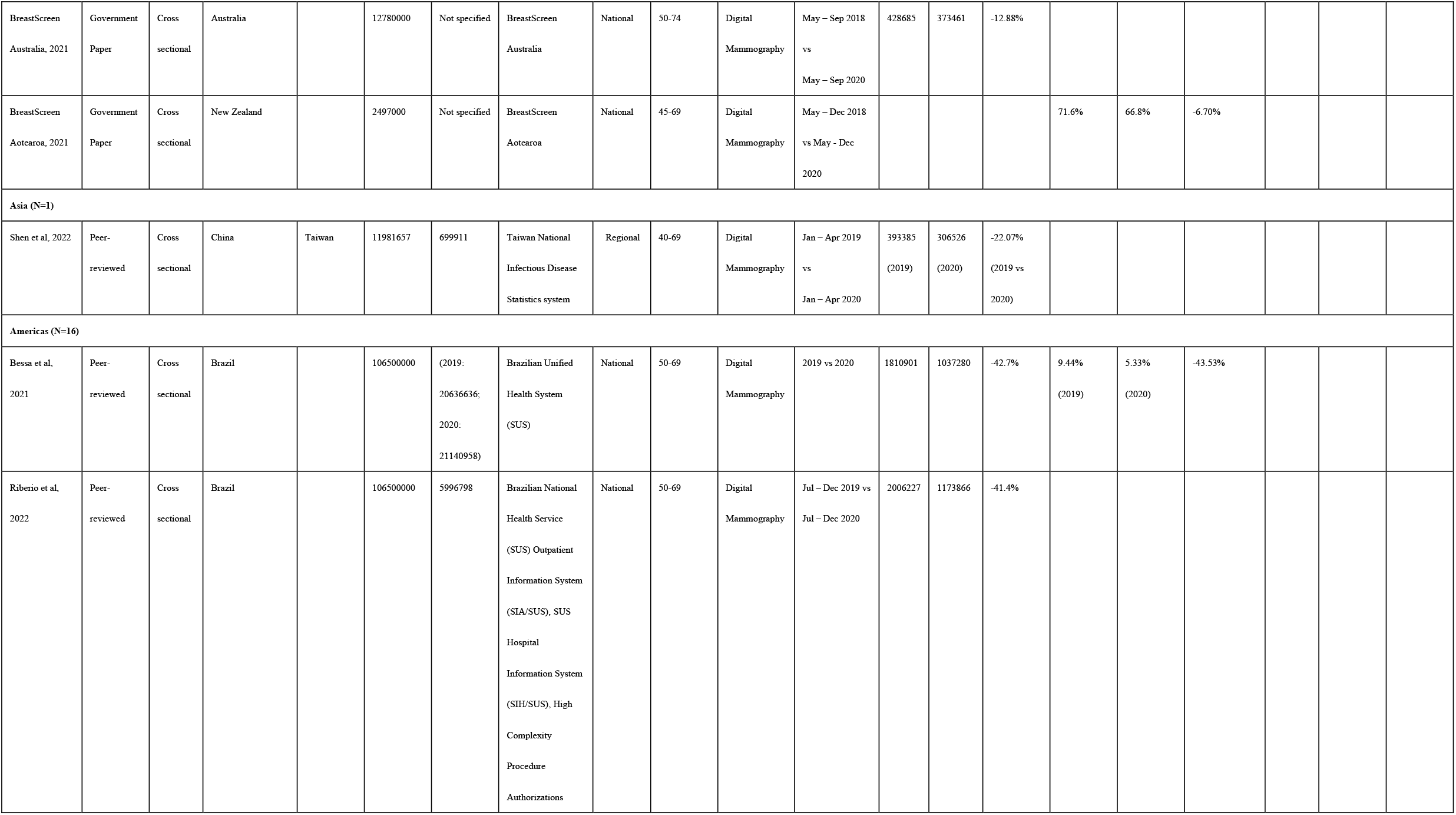
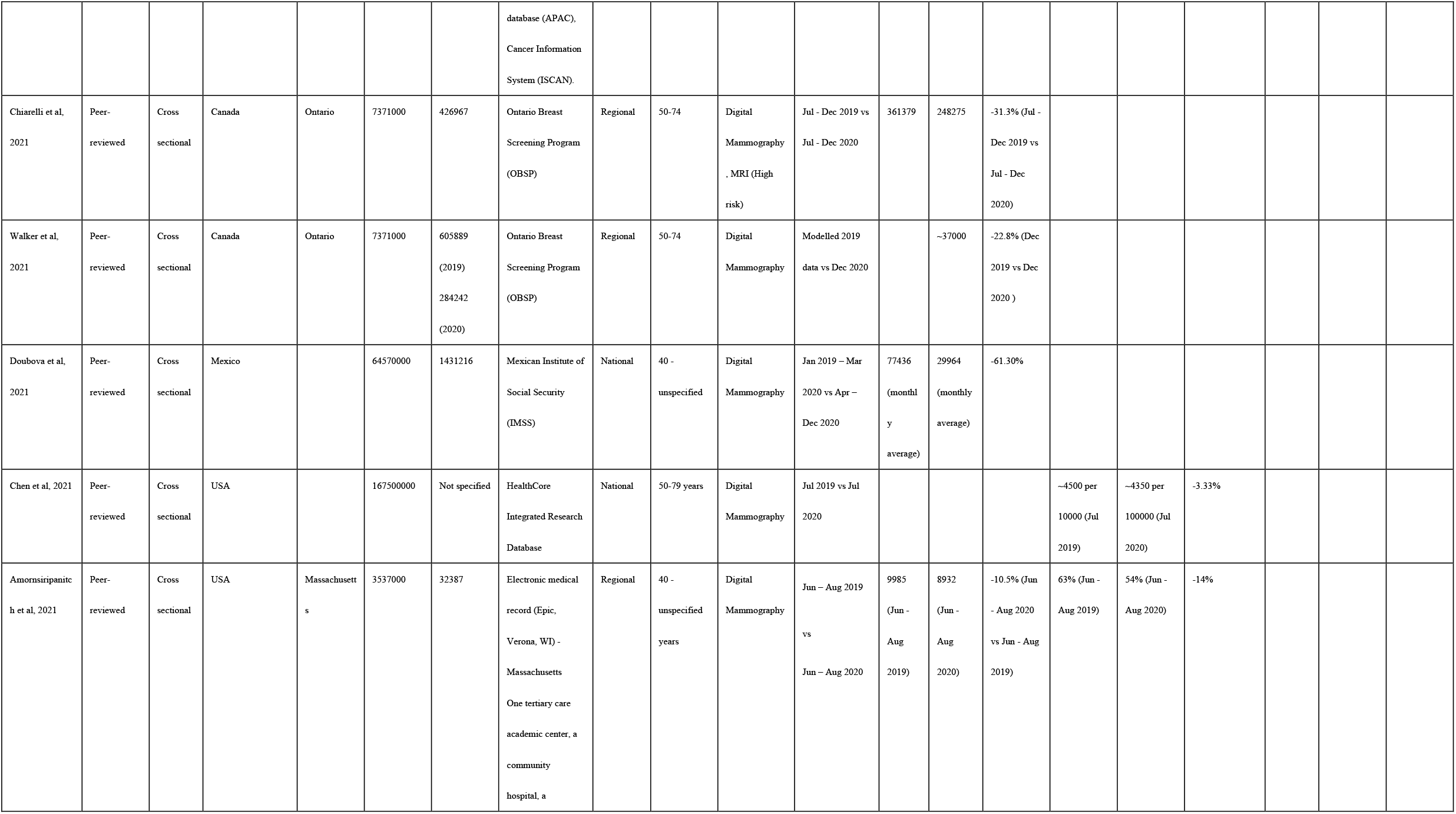
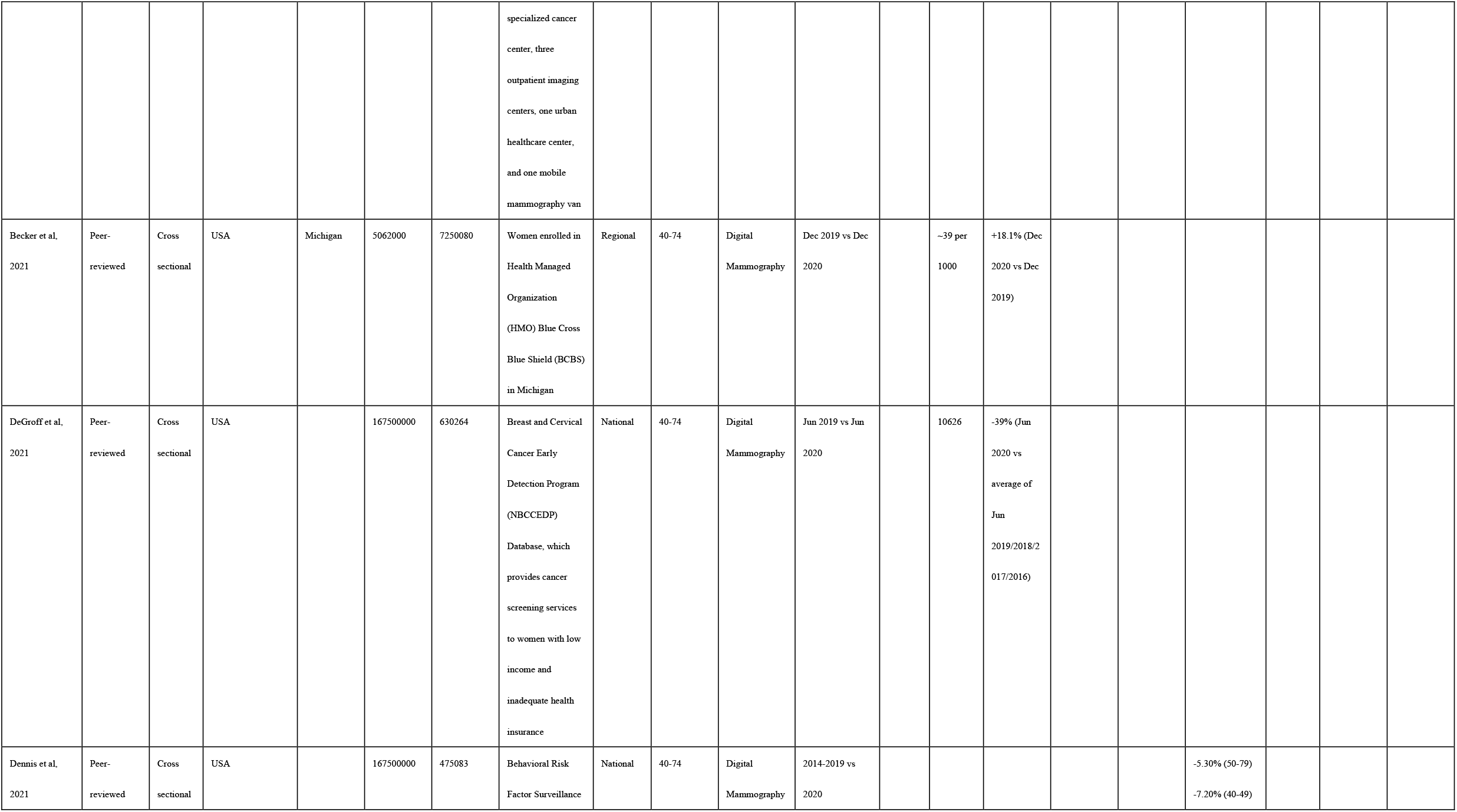
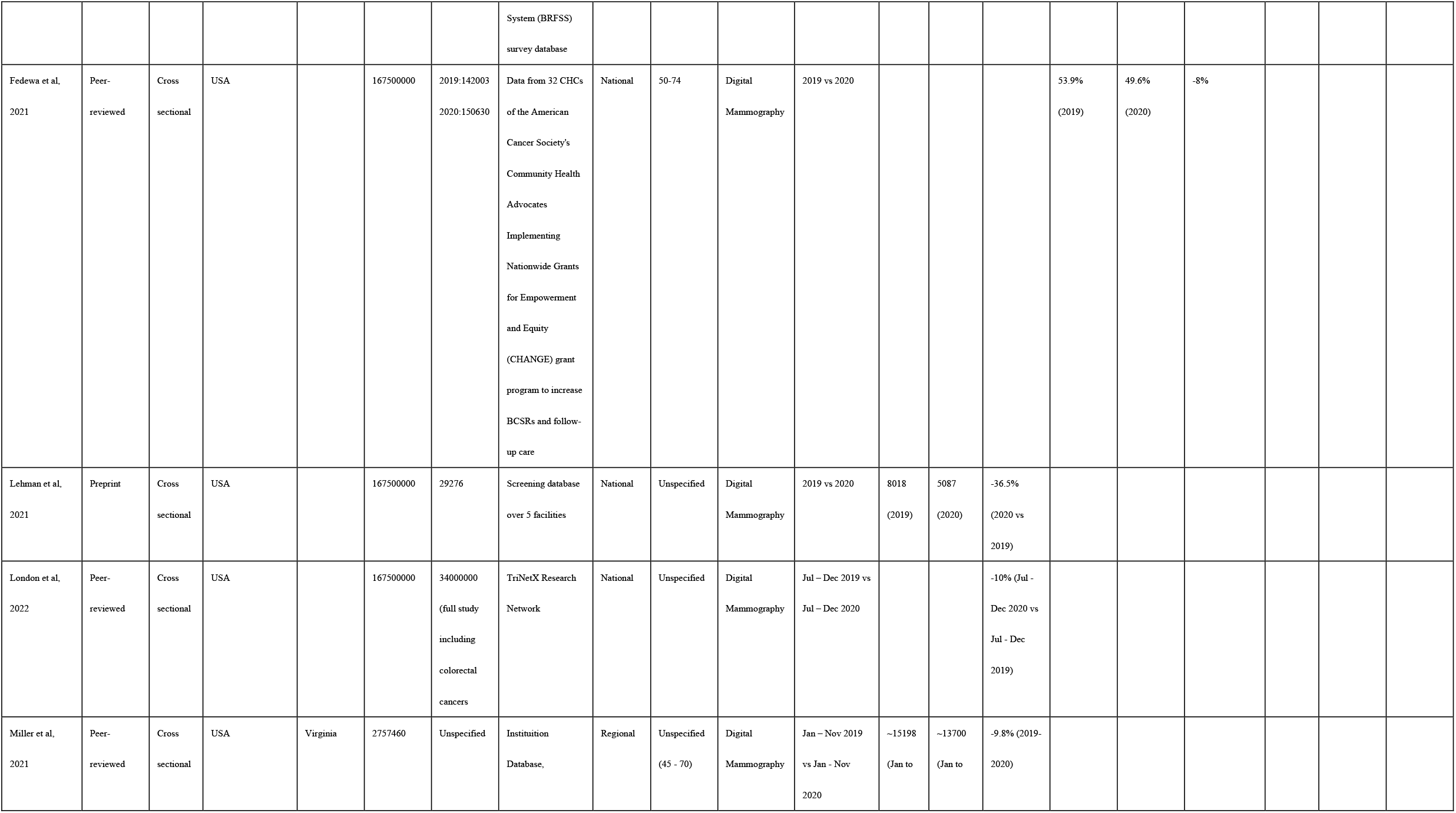
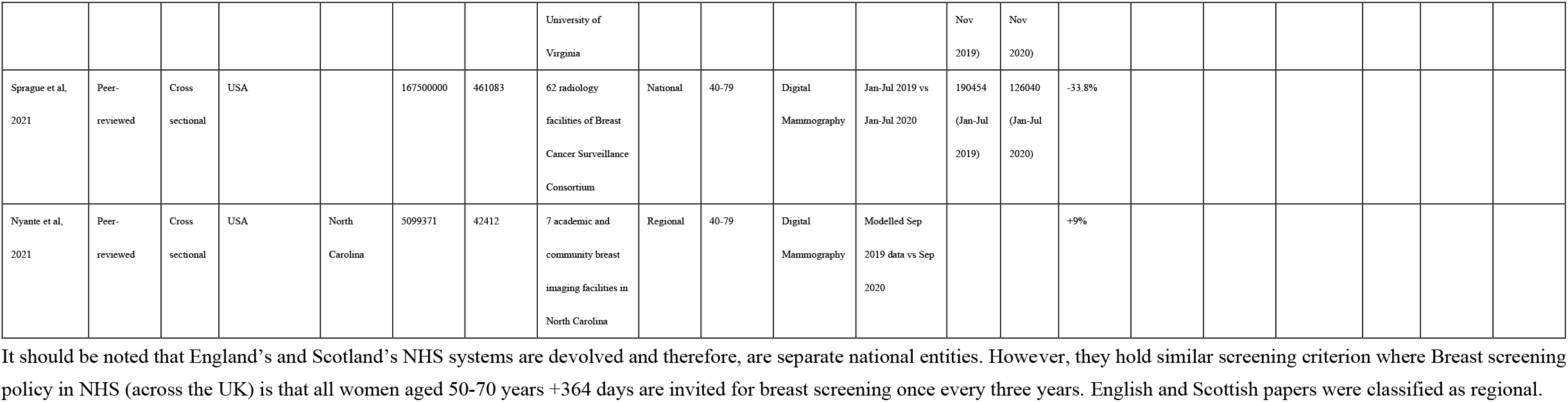
Descriptive characteristics of included cross-sectional studies (N=26)

| Study | Publication | Study | Country | Region (If | Total | Sample size | Study screening | Screening | Screening | Screening type | Screening time | Primary Outcome |  |  |  |  |  | Secondary outcome |  |  |
| --- | --- | --- | --- | --- | --- | --- | --- | --- | --- | --- | --- | --- | --- | --- | --- | --- | --- | --- | --- | --- |
|  | type | design |  | not | Female |  | data source | (National/<br>Regional) | age range |  | comparison | Volume | Volume | Volume | Participatio | Participatio | Participation | Incident | Incidence | Incidence |
|  |  |  |  |  |  |  |  |  |  |  |  | before | after | change | n rate | n rate after | rate change | e before | after | change |
|  |  |  |  |  | Population<br>of Study<br>Area |  |  |  |  |  |  | COVID | COVID |  | before | COVID |  | COVID | COVID |  |
| Europe (n=7) |  |  |  |  |  |  |  |  |  |  |  |  |  |  |  |  |  |  |  |  |
| Campbell et al,<br>2021 | Peer-reviewed | Cross<br>sectional | Scotland (UK) |  | 2728000 | Not<br>specified | NHS Scotland | Regional | 50-70 | Digital<br>Mammography | Aug – Dec 2019<br>vs Aug -Dec<br>2020 |  |  |  | 69% (Aug<br>2019)<br>73% (March<br>2020) | 80% (Aug<br>2020)<br>81% (Mar<br>2021) | +10.96% (Aug<br>2020 vs Aug<br>2019)<br>+2-8% (Sep –<br>Dec 2020 vs<br>Sep-Dec 2019) |  |  |  |
| Jidkova et al,<br>2022 | Peer-reviewed | Cross<br>sectional | Belgium | Flanders | 3382265 | Unspecified | Flanders Online<br>Screening Database | Regional | 50-69 | Digital<br>Mammography | Jul – Nov 2019<br>vs Jul – Nov<br>2020 |  |  |  |  |  | -1.0% (-1.3; -<br>0.7)- |  |  |  |
| Knoll et al, 2022 | Preprint | Cross<br>sectional | Austria | Innsbruck | 567300 | 596 | Database from<br>gynecological<br>oncological center<br>in Austria, Tyrol | Local | 45 - 69 years<br>invited for<br>screening.<br>Women aged<br>40 – 44 years<br>and 70 - 75<br>years may<br>opt in | Digital<br>Mammography | Mar – Dec 2019<br>vs Mar – Dec<br>2020 |  |  |  |  |  |  | 263 (Mar<br>- Dec<br>2019) | 212 (Mar -<br>Dec 2020) | -19.39% |
| Eijkelboom et al, 2021 | Peer-reviewed | Cross sectional | Netherlands |  | 8701000 | 3371 | Netherlands Cancer Registry | National | 50-75 | Digital Mammography | Jan – Feb 2020 vs Jul – Aug 2020 |  |  |  |  |  |  | 232.14 per week | 180.16 per week | -9.01% |
| Losurdo et al, 2022 | Peer-reviewed | Cross sectional | Italy | Friuli Venezia Giulia | 624418 | 58643 | “Data-Breast” database of the “Eusoma certified SSD Breast Unit of Trieste and from the Surgical Department of DAI Chirurgia Generale—ASUGL | Regional | 50-69 | Digital Mammography | Oct – Dec 2019 vs Oct – Dec 2020 |  |  | - 1.987<br>(– 11.9%)<br>(Oct - Dec 2020 vs 2019 FVG)<br>– 135.196<br>(– 23.7%)<br>(Oct - Dec 20 vs 2019 Italy) |  |  |  |  |  |  |
| Toss et al, 2021 | Peer-reviewed | Cross sectional | Italy | Northern Italy, Emilia Romagna | 2291000 | 24994 | Emilia Romagna National Healthcare System | Regional | 45-79 | Digital Mammography | 2019 vs 2020 |  |  |  |  |  |  | 223 out of 15942<br>0.01399<br>(2019) | 177 out of 9052<br>0.01955<br>(2020) | -39.74% |
| NHS England, 2021 | Government paper | Cross sectional | England (UK) |  | 33940000 | "2.23m<br>(2019) | NHS England | Regional | 50-71 | Digital Mammography | 2019 vs 2020 | 176666<br>(2019<br>per<br>month) | 148750<br>(2020 per<br>month) | -15.80%<br>(Monthly<br>Average<br>2019 vs<br>Monthly<br>Average<br>2020) | 69.1%<br>(2019) | 61.8%<br>(2020) | -11.80% |  |  | +8.30% |
| Oceania (n=2) |  |  |  |  |  |  |  |  |  |  |  |  |  |  |  |  |  |  |  |  |
| BreastScreen<br>Australia, 2021 | Government<br>Paper | Cross<br>sectional | Australia |  | 12780000 | Not specified | BreastScreen<br>Australia | National | 50-74 | Digital<br>Mammography | May – Sep 2018<br>vs<br>May – Sep 2020 | 428685 | 373461 | -12.88% |  |  |  |  |  |  |
| BreastScreen<br>Aotearoa, 2021 | Government<br>Paper | Cross<br>sectional | New Zealand |  | 2497000 | Not specified | BreastScreen<br>Aotearoa | National | 45-69 | Digital<br>Mammography | May – Dec 2018<br>vs May - Dec<br>2020 |  |  |  | 71.6% | 66.8% | -6.70% |  |  |  |
| Asia (N=1) |  |  |  |  |  |  |  |  |  |  |  |  |  |  |  |  |  |  |  |  |
| Shen et al, 2022 | Peer-reviewed | Cross<br>sectional | China | Taiwan | 11981657 | 699911 | Taiwan National<br>Infectious Disease<br>Statistics system | Regional | 40-69 | Digital<br>Mammography | Jan – Apr 2019<br>vs<br>Jan – Apr 2020 | 393385<br>(2019) | 306526<br>(2020) | -22.07%<br>(2019 vs<br>2020) |  |  |  |  |  |  |
| Americas (N=16) |  |  |  |  |  |  |  |  |  |  |  |  |  |  |  |  |  |  |  |  |
| Bessa et al,<br>2021 | Peer-reviewed | Cross<br>sectional | Brazil |  | 106500000 | (2019:<br>20636636;<br>2020:<br>21140958) | Brazilian Unified<br>Health System<br>(SUS) | National | 50-69 | Digital<br>Mammography | 2019 vs 2020 | 1810901 | 1037280 | -42.7% | 9.44%<br>(2019) | 5.33%<br>(2020) | -43.53% |  |  |  |
| Riberio et al,<br>2022 | Peer-reviewed | Cross<br>sectional | Brazil |  | 106500000 | 5996798 | Brazilian National<br>Health Service<br>(SUS) Outpatient<br>Information System<br>(SIA/SUS), SUS<br>Hospital<br>Information System<br>(SIH/SUS), High<br>Complexity<br>Procedure<br>Authorizations database (APAC),<br>Cancer Information<br>System (ISCAN). | National | 50-69 | Digital<br>Mammography | Jul – Dec 2019 vs<br>Jul – Dec 2020 | 2006227 | 1173866 | -41.4% |  |  |  |  |  |  |
| Chiarelli et al,<br>2021 | Peer-reviewed | Cross<br>sectional | Canada | Ontario | 7371000 | 426967 | Ontario Breast<br>Screening Program<br>(OBSP) | Regional | 50-74 | Digital<br>Mammography<br>, MRI (High<br>risk) | Jul - Dec 2019 vs<br>Jul - Dec 2020 | 361379 | 248275 | -31.3% (Jul -<br>Dec 2019 vs<br>Jul - Dec<br>2020) |  |  |  |  |  |  |
| Walker et al,<br>2021 | Peer-reviewed | Cross<br>sectional | Canada | Ontario | 7371000 | 605889<br>(2019)<br>284242<br>(2020) | Ontario Breast<br>Screening Program<br>(OBSP) | Regional | 50-74 | Digital<br>Mammography | Modelled 2019<br>data vs Dec 2020 |  | ~37000 | -22.8% (Dec<br>2019 vs Dec<br>2020 ) |  |  |  |  |  |  |
| Doubova et al,<br>2021 | Peer-reviewed | Cross<br>sectional | Mexico |  | 64570000 | 1431216 | Mexican Institute of<br>Social Security<br>(IMSS) | National | 40 -<br>unspecified | Digital<br>Mammography | Jan 2019 – Mar<br>2020 vs Apr –<br>Dec 2020 | 77436<br>(monthl<br>y<br>average) | 29964<br>(monthly<br>average) | -61.30% |  |  |  |  |  |  |
| Chen et al, 2021 | Peer-reviewed | Cross<br>sectional | USA |  | 167500000 | Not specified | HealthCore<br>Integrated Research<br>Database | National | 50-79 years | Digital<br>Mammography | Jul 2019 vs Jul<br>2020 |  |  |  | ~4500 per<br>10000 (Jul<br>2019) | ~4350 per<br>100000 (Jul<br>2020) | -3.33% |  |  |  |
| Amornsiripanitch et al, 2021 | Peer-reviewed | Cross<br>sectional | USA | Massachusetts | 3537000 | 32387 | Electronic medical<br>record (Epic,<br>Verona, WI) -<br>Massachusetts<br>One tertiary care<br>academic center, a<br>community<br>hospital, a | Regional | 40 -<br>unspecified<br>years | Digital<br>Mammography | Jun – Aug 2019<br>vs<br>Jun – Aug 2020 | 9985<br>(Jun -<br>Aug<br>2019) | 8932<br>(Jun -<br>Aug<br>2020) | -10.5% (Jun<br>- Aug 2020<br>vs Jun - Aug<br>2019) | 63% (Jun -<br>Aug 2019) | 54% (Jun -<br>Aug 2020) | -14% |  |  |  |

|  |  |  |  |  |  |  |  |  |  |  |  |  |  |  |  |  |  |
| --- | --- | --- | --- | --- | --- | --- | --- | --- | --- | --- | --- | --- | --- | --- | --- | --- | --- |
|  |  |  |  |  |  |  | specialized cancer center, three outpatient imaging centers, one urban healthcare center, and one mobile mammography van |  |  |  |  |  |  |  |  |  |  |
| Becker et al, 2021 | Peer-reviewed | Cross sectional | USA | Michigan | 5062000 | 7250080 | Women enrolled in Health Managed Organization (HMO) Blue Cross Blue Shield (BCBS) in Michigan | Regional | 40-74 | Digital Mammography | Dec 2019 vs Dec 2020 |  | ~39 per 1000 | +18.1% (Dec 2020 vs Dec 2019) |  |  |  |
| DeGroff et al, 2021 | Peer-reviewed | Cross sectional | USA |  | 167500000 | 630264 | Breast and Cervical Cancer Early Detection Program (NBCCEDP) Database, which provides cancer screening services to women with low income and inadequate health insurance | National | 40-74 | Digital Mammography | Jun 2019 vs Jun 2020 |  | 10626 | -39% (Jun 2020 vs average of Jun 2019/2018/2017/2016) |  |  |  |
| Dennis et al, 2021 | Peer-reviewed | Cross sectional | USA |  | 167500000 | 475083 | Behavioral Risk Factor Surveillance | National | 40-74 | Digital Mammography | 2014-2019 vs 2020 |  |  |  |  |  | -5.30% (50-79)<br>-7.20% (40-49) |

|  |  |  |  |  |  |  |  |  |  |  |  |  |  |  |  |  |  |
| --- | --- | --- | --- | --- | --- | --- | --- | --- | --- | --- | --- | --- | --- | --- | --- | --- | --- |
|  |  |  |  |  |  |  | System (BRFSS)<br>survey database |  |  |  |  |  |  |  |  |  |  |
| Fedewa et al,<br>2021 | Peer-reviewed | Cross<br>sectional | USA |  | 167500000 | 2019:142003<br>2020:150630 | Data from 32 CHCs<br>of the American<br>Cancer Society's<br>Community Health<br>Advocates<br>Implementing<br>Nationwide Grants<br>for Empowerment<br>and Equity<br>(CHANGE) grant<br>program to increase<br>BCSRs and follow-up care | National | 50-74 | Digital<br>Mammography | 2019 vs 2020 |  |  |  | 53.9%<br>(2019) | 49.6%<br>(2020) | -8% |
| Lehman et al,<br>2021 | Preprint | Cross<br>sectional | USA |  | 167500000 | 29276 | Screening database<br>over 5 facilities | National | Unspecified | Digital<br>Mammography | 2019 vs 2020 | 8018<br>(2019) | 5087<br>(2020) | -36.5%<br>(2020 vs<br>2019) |  |  |  |
| London et al,<br>2022 | Peer-reviewed | Cross<br>sectional | USA |  | 167500000 | 34000000<br>(full study<br>including<br>colorectal<br>cancers | TriNetX Research<br>Network | National | Unspecified | Digital<br>Mammography | Jul – Dec 2019 vs<br>Jul – Dec 2020 |  |  | -10% (Jul -<br>Dec 2020 vs<br>Jul - Dec<br>2019) |  |  |  |
| Miller et al,<br>2021 | Peer-reviewed | Cross<br>sectional | USA | Virginia | 2757460 | Unspecified | Institution<br>Database, | Regional | Unspecified<br>(45 - 70) | Digital<br>Mammography | Jan – Nov 2019<br>vs Jan - Nov<br>2020 | ~15198<br>(Jan to | ~13700<br>(Jan to | -9.8% (2019-2020) |  |  |  |
|  |  |  |  |  |  |  | University of Virginia |  |  |  |  | Nov 2019) | Nov 2020) |  |  |  |  |
| Sprague et al, 2021 | Peer-reviewed | Cross sectional | USA |  | 167500000 | 461083 | 62 radiology facilities of Breast Cancer Surveillance Consortium | National | 40-79 | Digital Mammography | Jan-Jul 2019 vs Jan-Jul 2020 | 190454 (Jan-Jul 2019) | 126040 (Jan-Jul 2020) | -33.8% |  |  |  |
| Nyante et al, 2021 | Peer-reviewed | Cross sectional | USA | North Carolina | 5099371 | 42412 | 7 academic and community breast imaging facilities in North Carolina | Regional | 40-79 | Digital Mammography | Modelled Sep 2019 data vs Sep 2020 |  |  | +9% |  |  |  |

It should be noted that England's and Scotland's NHS systems are devolved and therefore, are separate national entities. However, they hold similar screening criterion where Breast screening policy in NHS (across the UK) is that all women aged 50-70 years +364 days are invited for breast screening once every three years. English and Scottish papers were classified as regional.

**Table 3.** Breast cancer screening volumes change among 106,484,908 subjects from eight countries.

| Change in Volume of Breast Cancer Screening (N=17) |  |  |  |  |  |  |
| --- | --- | --- | --- | --- | --- | --- |
| Study | Country | Region | National/Regional | Sample size | Screening timeframe comparison | Volume change |
| Europe (n=2) |  |  |  |  |  |  |
| Losurdo *et al.* 2022 | Italy | Friuli Venezia Giulia | Regional | 58643 | Oct – Dec 2019 vs Oct – Dec 2020 | -23.70% |
| NHS England, 2021 | UK | England | Regional | 3420000 | Monthly Average 2019 vs Monthly Average 2020 | -15.80% |
| Oceania (n=1) |  |  |  |  |  |  |
| BreastScreen Australia, 2021 | Australia | NA | National | 802146 | May – Sep 2018 vs May – Sep 2020 | -12.86% |
| Asia (n=1) |  |  |  |  |  |  |
| Shen *et al.* 2022 | China | Taiwan | Regional | 699911 | Jan – Apr 2019 vs Jan – Apr 2020 | -22.07% |
| America (n=13) |  |  |  |  |  |  |
| Bessa *et al.* 2021 | Brazil | NA | National | (2019: 20636636; 2020: 21140958) | 2019 vs 2020 | -42.72% |
| Riberio *et al.* 2022 | Brazil | NA | National | 5996798 | Jul – Dec 2019 vs Jul – Dec 2020 | -41.49% |
| Doubova *et al.* 2021 | Mexico | NA | National | 1431216 | Jan 2019 – Mar 2020 vs Apr – Dec 2020 | -61.30% |
| Chiarelli *et al.* 2021 | Canada | Ontario | Regional | 426967 | Jul - Dec 2019 vs Jul - Dec 2020 | -31.30% |
| Walker *et al.* 2021 | Canada | Ontario | Regional | 890131 | Modelled 2019 data vs Dec 2020 | -22.80% |
| Lehman *et al.* 2021 | USA | NA | National | 29276 | 2019 vs 2020 | -36.50% |
| Miller *et al.* 2021 | USA | North Carolina | Regional | 8536000 | Jan – Nov 2019 vs Jan – Nov 2020 | -9.80% |
| Amornsiripanitch *et al.* 2021 | USA | Massachusetts | Regional | 32387 | Jun – Aug 2019 vs Jun – Aug 2020 | -10.50% |
| London *et al.* 2022 | USA | NA | National | 34000000 | Jul – Dec 2019 vs Jul – Dec 2020 | -10.00% - +30.00% |
| DeGroff *et al.* 2021 | USA | NA | National | 630264 | Jun 2019 vs Jun 2020 | -39.00% |
| Becker *et al.* 2021 | USA | Michigan | Regional | 7250080 | Dec 2019 vs Dec 2020 | 18.10% |
| Sprague *et al.* 2021 | USA | NA | National | 461083 | Jul 2019 vs Jul 2020 | -10.30% (-20.40%-6.60%) |
| Nyante *et al.* 2021 | USA | North Carolina | Regional | 42412 | Modelled Sep 2019 data vs Sep 2020 | 9.00% (-12.00%-32.00%) |
| NA: Not applicable |  |  |  |  |  |  |
| Colour Legend (Breast Screening programme of study population): Green = Population-based Screening present in country; Yellow = Population-based screening present in country but private sector databases included (Brazilian National Health Service (SUS), Outpatient Information System (SIA/SUS), SUS Hospital Information System (SIH/SUS), High Complexity Procedure Authorizations database (APAC), Cancer Information System (ISCAN)); Red = Privatised system with mix of national and private insurance usage; Blue = The National Breast and Cervical Cancer Early Detection Program (NBCCEDP) that provides cancer screening services to women with low income and inadequate health insurance.; Grey = Health Managed Care Organization (HMO) -based screening [database covers HMO data from Michigan]. For studies conducted in the USA, ACS guidelines were used as the data collection comparator starting point where Mar-Jun 2020 was considered to be a suspension in screening. |  |  |  |  |  |  |

**Table 4.** Breast cancer screening participation rates change from nine studies from five countries.

| Change in Participation Uptake rate of Breast Cancer Screening (N=9) |  |  |  |  |  |  |
| --- | --- | --- | --- | --- | --- | --- |
| Study | Country | Region | National/Regional | Sample size | Screening timeframe comparison | Participation rate change |
| Europe (n=3) |  |  |  |  |  |  |
| NHS England, 2021 | UK | England | Regional | 3420000 | 2019 vs 2020 | -11.80% |
| Campbell *et al.* 2021 | UK | Scotland | Regional | NA | Aug – Dec 2019 vs Aug – Dec 2020 | +10.96% (Aug 2020)<br>+2-8% (Sep – Dec 2020) |
| Jidkova *et al.* 2022 | Belgium | Flanders | Regional | NA | Jul – Dec 2019 vs Jul – Dec 2020 | -1.0% (-1.3; -0.7) |
| Oceania (n=1) |  |  |  |  |  |  |
| BreastScreen Aotearoa. 2021 | New Zealand | NA | National | NA | Dec 2018/2019 vs May – Dec 2020 | -6.70% |
| Americas (n=5) |  |  |  |  |  |  |
| Bessa *et al.* 2021 | Brazil | NA | National | (2019: 20636636;<br>2020: 21140958) | 2019 vs 2020 | -43.54% |
| Dennis *et al.* 2021 | USA | NA | National | 475083 (Age: 50-74)<br>117498 (Age: 40-49) | 2014-2019 vs 2020 | -5.30% (50-79)<br>-7.20% (40-49) |
| Fedewa *et al.* 2021 | USA | NA | National | 434840 | 2019 vs 2020 | -8.00% |
| Amornsiripanitch *et al.* 2021 | USA | Massachusetts | Regional | 32387 | Jun – Aug 2019 vs Jun – Aug 2020 | -14.80% |
| Chen *et al.* 2021 | USA | NA | National | NA | Jul 2019 vs Jul 2020 | -3.33% |
| NA: Not applicable |  |  |  |  |  |  |
| Colour Legend (Breast Screening programme of study population): Green = Population-based Screening available in country; Red = Privatised system with mix of national and private insurance usage; For studies conducted in the USA, ACS guidelines were used as the data collection comparator starting point where Mar-Jun 2020 was considered to be a suspension in screening |  |  |  |  |  |  |

### Data Synthesis

Data were synthesized descriptively since a meta-analysis was not appropriate due to heterogeneity of data. Data was collected by comparing outcome measures before and after COVID-19 infection control measures were introduced; this was presumed due to the worldwide prevalence of COVID-19 by March 2020. This included changes in breast screening volume, participation rates and breast cancer diagnosis rates (Figueroa *et al, 2021;* WHO, 2020).

Data were obtained from any point after lifting of COVID-19 breast screening suspension measures until an endpoint of 31 December 2020. If quantitative data was limited, the last data point of the study was analyzed. This was compared to data from an analogous pre-COVID-19 period in 2018-2019, or if data was unavailable, against any relevant pre-pandemic period. For countries with no breast screening suspension in 2020, data from during the COVID-19 pandemic was compared with an analogous period of 2018-2019. This phenomenon only occurred in Taiwan, China (Shen et al. 2022). A percentage change against the overall comparator period was calculated.

## Results

**Figure 1** summarizes the search strategy. The initial search retrieved 1207 articles and 935 independent records. After screening (see Methods), 26 cross-sectional studies from 12 countries were eligible for inclusion (**Table 2)**. Seven reports came from Europe (Campbell et al, 2021; Jidkova et al, 2022; Knoll et al, 2022; Eijkelboom et al, 2021; Losurdo et al, 2022; Toss et al, 2021; NHS England, 2021), two from Oceania (BreastScreen Australia, 2021; BreastScreen Aoteroa, 2021), one from Asia (Shen et al, 2022), two from South America (Bessa et al, 2021; Riberio et al, 2022) and 14 from North America (Chiarelli et al, 2021; Walker et al, 2021; Doubouva et al, 2021; Chen et al, 2021; Amornsiripanitch et al, 2021; Becker et al, 2021; DeGroff et al, 2021; Dennis et al, 2021; Fedewa et al, 2021; Lehman et al, 2021; London et al, 2022; Miller et al, 2021; Sprague et al, 2021; Nyante et al, 2021). The most frequently reported country was the USA (n = 11). Studies examined either regional (n = 13) or national populations (n = 13). Analysis of data from all studies was limited from 1 January 2020 to 31 December 2020.

**Figure 1:**
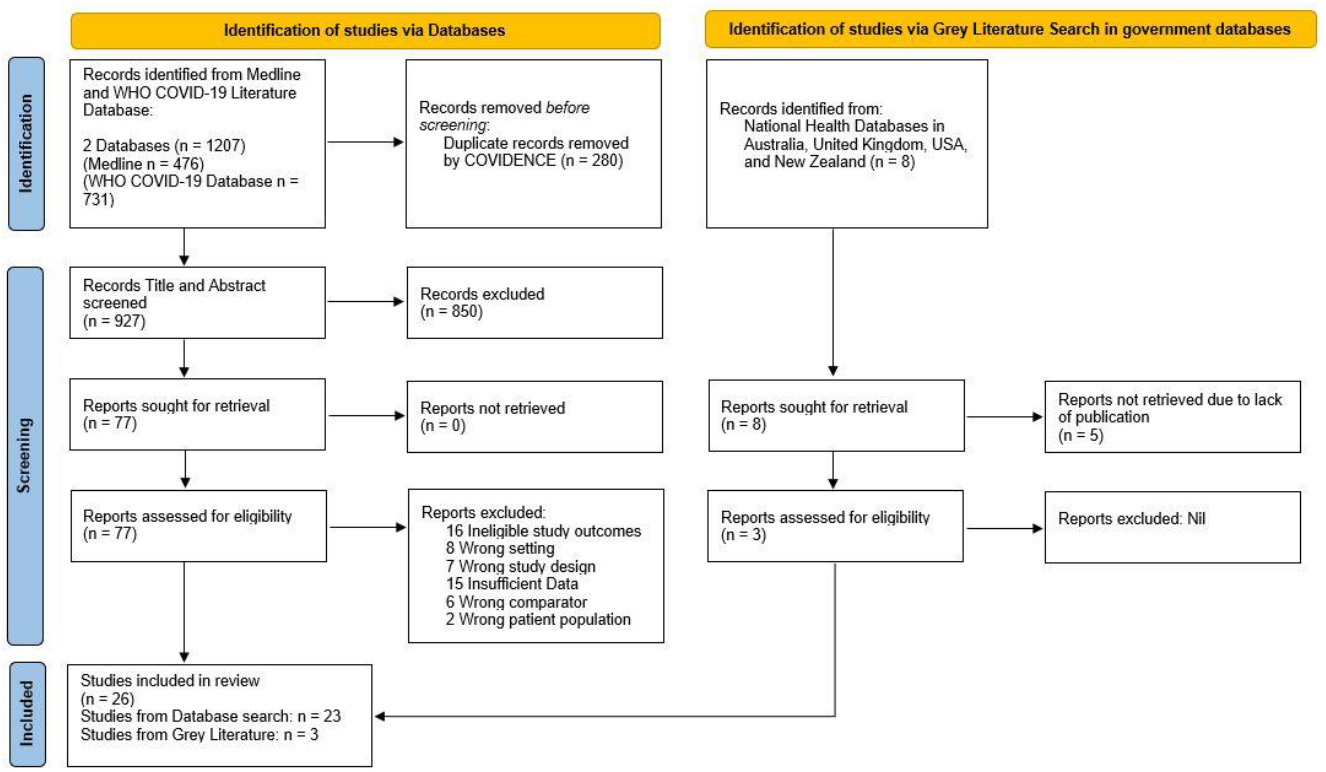
PRISMA diagram for showing screening process (adapted from Page et al. 2021)

### Screening volume changes over study period

Summary data from 17 studies in eight countries reporting breast cancer screening volumes, data from 106,484,908 women before and after COVID-19 infection control measures were extracted (data from 2017 to 2020 were the comparison time period, **Table 3;** Doubouva et al, 2021; Bessa et al, 2021; Riberio et al, 2022; Chiarelli et al, 2021; Losurdo et al, 2022; Walker et al, 2021; NHS England, 2021; Shen et al, 2022; BreastScreen Australia, 2021; DeGroff et al, 2021; Lehman et al, 2021; Amornsiripantich et al, 2021; Sprague et al, 2021; London et al, 2022; Miller et al, 2021; Nyante et al, 2021; Becker et al, 2021). Most studies that showed calendar period trends of screening volume, noted temporal variation with declines especially at the height of the pandemic between March-May 2020. In countries with national screening programs, a negative change in screening volume was reported with the lowest volume change estimated at - 12.86% in Australia (BreastScreen Australia, 2021) followed by -31.30% in Ontario, Canada (Chiarelli et al, 2021). A larger negative change in screening volume was observed in Brazil (−41.49%) (Ribero et al, 2022) and Mexico (−61.30%) (Doubouva et al, 2021. It should be noted that Brazil and Mexico have a lower proportion of population-based breast screening coverage relative to other countries; Brazil having coverage of ∼24%, and Mexico having ∼20% coverage of the eligible population (OECD, 2021; Unger-Saldaña et al, 2020). A significant proportion of breast screening in Brazil and Mexico consists of opportunistic screening programs.

In the USA, which has mix of insurance providers there was a wide range of change in screening volume. Using data from Health Managed Organization (HMO) Blue Cross Blue Shield (BCBS) from the state of Michigan, the authors observed temporal changes in rates with an increase slightly above 2019 levels in the last few months of the year, with an 18.10% overall increase in screening volume (Becker et al. 2021). However, it’s important to note that although rates were above 2019 levels, the authors noted that the odds that a woman received breast cancer screening remained 20% lower in 2020 relative to 2019 (Becker et al. 2021). This was consistent with the decrease in screening volume that was generally observed from six studies with data among populations wholly or partially covered by national insurance (Lehman et al, 2021; Amornsiripanitch et al, 2021; Sprague et al, 2021; London et al, 2022; Miller et al, 2021; Nyante et al, 2021). Percentage decreases ranged from -36.50% (Lehman et al. 2021) to -9.80% (Miller et al. 2021). Data from the USA National Breast and Cervical Cancer Early Detection Program (NBCCEDP), which provides cancer screening services to women with low income and inadequate health insurance, reported a greater decrease (−39.00%) in volume (DeGroff et al. 2021). Two other studies had smaller populations with less certainty and wider confidence intervals with one reporting an 8% increase (Nyante et al. 2021) and the other a -10% decline (London et al. 2021). In the USA, where there is a mix of national (Medicare) and private insurance depending on age, screening volume changes were similar to other national screening programs at -36.50% (Lehman et al, 2021). In contrast, a positive increase in volume was observed among private insurance providers +30% (London et al, 2022)

### Screening participation rate changes

A total of nine cross-sectional studies reported breast cancer screening participation rates and represented > 46,257,402 participants from varying calendar periods across five countries (Amornsiripantich et al, 2021; Dennis et al, 2021; Fedewa et al, 2021; Chen et al, 2021; NHS England, 2021; Campbell et al, 2021; Bessa et al, 2021; BreastScreen Aoteroa, 2021; Jidkova et al, 2022). There was considerable variability in change (**Table 4**), ranging from +2-8% in Scotland to -43.54% in Brazil (Campbell et al, 2021; Bessa et al, 2021). In the USA, with a mix of national and private insurance usage, there was a consistent negative change in screening participation rates (Amornsiripantich et al, 2021; Dennis et al, 2021; Fedewa et al, 2021; Chen et al, 2021).

### Study quality

The quality of included studies was assessed using the JBI tool (**Table 5**). A weakness across most studies was failure to identify and consider confounding factors. From **Table 5**, twenty-five studies had no issues defining the inclusion sample. Nineteen studies were clear in defining the study setting and subjects. Studies had no issues quantifying exposure of COVID-19, although this was based on temporality since all healthcare systems globally were affected (Worldometer, 2022). All studies apart from Becker et al (2021) had no issue measuring the condition through either screening appointment attendance or insurance claims data. Most studies (65%, N=17) did not define confounding factors regarding measurement of primary outcomes. Regarding comparison of volumes of screening prior to COVID-19 and observed periods, these studies did not provide source of reduction in screening capacity (e.g. due to social distancing or participation uptake). Twenty-three studies failed to provide strategies to address confounding factors (e.g., elucidating reduction in capacity and presenting it as a proportion to overall volume).

**Table 5.** Summary of results of appraisal of all included studies with JBI Critical appraisal tool for Cross-sectional Studies.

| Joanna Briggs Institute Critical Appraisal Tool for Cross-Sectional Studies Appraisal Table |  |  |  |  |  |  |  |  |
| --- | --- | --- | --- | --- | --- | --- | --- | --- |
| Study | 1. Were the criteria for inclusion in the sample clearly defined? | 2. Were the study subjects and the setting described in detail? | 3. Was the exposure measured in a valid and reliable way? | 4. Were objective, standard criteria used for measurement of the condition? | 4. Were confounding factors identified? | 5. Were strategies to deal with confounding factors stated? | 7. Were the outcomes measured in a valid and reliable way? | 8. Was appropriate statistical analysis used? |
| Amomsiripanitch *et al.* 2021 | Y | Y | Y | Y | Y | N | Unclear | Unclear |
| Becker *et al.* 2021 | Y | Y | Y | N | Y | N | N | Unclear |
| Bessa *et al.* 2021 | Y | Unclear | Y | Y | N | N | N | Unclear |
| Campbell *et al.* 2021 | Y | Unclear | Y | Y | Unclear | N | Y | Unclear |
| Chen *et al.* 2021 | Y | Unclear | Y | Y | Y | N | Y | Unclear |
| Chiarelli *et al.* 2021 | Y | Y | Y | Y | Unclear | N | Y | Unclear |
| DeGroff *et al.* 2021 | Y | Y | Y | Y | Y | N | Y | Unclear |
| Dennis *et al.* 2021 | Y | Y | Y | Y | N | N | Y | Unclear |
| Dobova *et al.* 2021 | Y | Unclear | Y | Y | N | N | N | Y |
| Jidkova *et al.* 2022 | Y | Y | Y | Y | Unclear | N | Y | Unclear |
| Knoll *et al.* 2022 | Y | Y | Y | Y | N | N | Y | Unclear |
| Fedewa *et al.* 2021 | Y | Y | Y | Y | N | N | Y | Unclear |
| BreastScreen Australia. 2021 | Y | Y | Y | Y | N | N | Y | N |
| Eijkelboom *et al.* 2021 | Y | Y | Y | Y | Y | Y | Y | Unclear |
| Lehman *et al.* 2021 | N | N | Y | Y | Y | N | Y | Unclear |
| London *et al.* 2022 | N | N | Y | Y | N | N | N | Unclear |
| Losurdo *et al.* 2022 | Y | Y | Y | Y | N | N | Y | Unclear |
| Walker *et al.* 2021 | Y | Y | Y | Y | Unclear | N | Y | Unclear |
| Toss *et al.* 2021 | Y | Y | Y | Y | N | N | Y | Unclear |
| Shen *et al.* 2022 | Y | Y | Y | Y | Unclear | N | Y | Unclear |
| Riberio *et al.* 2022 | Y | Y | Y | Y | N | N | Y | Unclear |
| Miller *et al.* 2021 | Y | Unclear | Y | Y | N | N | Y | Y |
| Sprague *et al.* 2021 | Y | Y | Y | Y | Unclear | Y | Y | Y |
| Nyante *et al.* 2021 | Y | Y | Y | Y | Y | Y | Y | Y |
| NHS England, 2021 | Y | Y | Y | Y | N | N | Y | N |
| BreastScreen Aotearoa. 2021 | Y | Y | Y | Y | N | N | Y | N |
| Colour Legend: Green = Yes; Yellow = Unclear; Orange = No |  |  |  |  |  |  |  |  |

Four studies (Bessa et al. 2021; Becker et al. 2021; London et al. 2022; Doubova et al. 2021) had unclear reasons for selection of study subjects and control groups (London et al, 2022), confounding factors that were not indicated, nor strategies included to solve this. Among these four papers, vague definition of control groups resulted in a poor comparator, resulting in unreliable outcome measures.

Twenty-three studies provided basic statistical analyses (e.g. mean, adjusted rates per population) with basic data presentation. Statistical analyses were not performed in three government papers (BreastScreen Australia. 2021; NHS England. 2021; BreastScreen Aotearoa. 2021). Twenty-two studies were unclear or did not provide sufficient descriptive statistical analyses regarding comparison of control data to observed data. Statistical analyses were performed in four studies. This includes provision of odds ratios by Doubova et al (2021) and Miller et al (2021), Poisson estimation of a 95% confidence interval by Sprague et al (2021) and 95% confidence intervals from comparison of means from Nyante et al (2021).

## Discussion

In this rapid review we show that during the COVID-19 pandemic, there was a generally reported reduction in breast cancer screening volume and participation that varied by healthcare setting (e.g., national population-based screening vs opportunistic or private health care). Our data suggests that participation is a major contributor, which requires monitoring by health systems and could inform prevention and early diagnosis efforts, especially if certain groups are not participating.

Fear of COVID-19 infection is likely an important source that would reduce screening participation and volumes. In populations with national population-based screening, there were competing risks between contracting COVID-19 and breast screening health benefits (Chiarelli et al. 2021; Walker et al. 2021). Considering initial lack of free access to vaccines in 2020, severity of the first COVID-19 wave, and older age of breast screening patients, patients likely translated to decreased screening participation due to infection fears (Losurdo et al. 2022). Governmental databases in England and New Zealand (NHS England, 2021; BreastScreen Aotearoa, 2021) corroborate this; participation uptake rates decreased by - 6.70% (New Zealand) to -11.80% (England). Given that uptake rate measures the proportion of patients who fulfil their appointment, monitoring this outcome could provide early indications on reductions in willingness of patients to attend screening. Shen et al (2022) describes changes in patient behaviour in Taiwan where they shifted mammography appointments from hospitals to mobile screening centres where perceived risks of contracting COVID-19 were lower. This led to greater decreases in hospital-based mammography volumes where a -41.43% reduction was observed relative to a -23.99% reduction in mobile centres. Interestingly, this phenomenon occurred despite low COVID-19 prevalence in Taiwan, China.

Reductions in screening capacity are likely another source for screening volume reductions. Screening capacity reductions were caused by social distancing, staggered appointments, staff exposure to COVID-19, and cleaning measures. This likely resulted in reductions in time allocated for screening to occur (Walker et al. 2021; Sprague et al. 2020). Sprague et al (2021) considered screening capacity when assessing screening volume. Even though screening capacity recovered to pre-pandemic levels in July 2020, screening volume experienced a 10.8% decrease relative to the control period. Reductions in screening capacity were potentially not the sole factor to screening volume reductions. However, most publications included in our rapid review did not collect data regarding screening capacity so we cannot determine the proportion of change in screening volume that was attributed to either reduction in screening capacity or change in patient willingness to attend screening. Future analyses are needed where both of these measures are obtained, which would inform what measures are needed (e.g. information campaigns to alleviate patient fears or increase clinical staffing for catch-up of missed appointments).

Our data supports differences by healthcare system which were particularly evident in data from the USA where there is a mix of private and national healthcare (Medicare for persons 65+ (Medicare.gov). DeGroff et al (2021), who studied populations reliant solely on national health insurance, showed larger screening volume reductions (−39.00%). This was relative to studies focusing solely on populations with private insurances, or studies including patients from both groups (−36.50% to +30%). Amornsiripanitch et al (2021), which included national and private insurance patients, corroborates this. Medicaid and Medicare patients had - 17.06% screening volume reduction compared to -10.50% experienced by the entire population. Miller et al (2021) suggests opportunity-cost of attending breast screening in lower income groups (e.g., employment), may have led to decreased breast screening in such populations. Some literature showed increases in screening volumes (Nyante et al. 2021; Becker et al. 2021) and uptake rates (Campbell et al. 2021). Increased volume (+9% 95% CI [-12, 32]) from Nyante et al (2021) was inconclusive due to wide 95% confidence intervals. Although this study was robust limited data collection till September 2020 did not show full extent of change regarding screening volumes after lifting of COVID-19 suspension guidelines in June 2020. From trends explored in study, breast screening rates were possibly recovering in the study population (USA) in late-2020, but more data is required. The Affordable Care Act may have alleviated breast screening cost through health insurance coverage reforms (Zhao et al. 2020). However, this does not address other underlying socio-economic inequalities (e.g. high cost of treatment, time off from work due to sickness).

Patients from deprived backgrounds may be fearful of dealing with the consequences of abnormal screening results (e.g., treatment). This may strain patient finances worsened by COVID-19, potentially explaining lower screening volumes and uptake. Future data on patient characteristics including insurance status, socioeconomic and race/ethnicity could inform targeted campaigns to reduce inequities if disparities exist.

Becker et al (2021) showed a screening volume increase after lifting of COVID-19 suspension guidelines. This study focused on patients who utilize solely private insurance. Patients already paying for services may be more inclined to maximize utilization of coverage. However, this study states that the odds that a woman received breast cancer screening remained 20% lower in 2020 (OR = 0.80 (0.80, 0.81)) relative to 2019. This study scored poorly in the JBI appraisal tool due to poor outcome measurement; it was unclear how odds ratio was derived, therefore, increasing the risk of bias of this study. Unusual outcome measures were used, that being the claims invoice for the service. This appeared unreliable; it was unclear whether paying for the service equates to a fulfilled appointment. Invoices could be delayed, making it unclear when the screening took place. This study’s evidence quality needs to be increased for results to be conclusive.

Campbell et al (2021) states a 10.96% increase in uptake rate in Scotland. This study population (within study period) solely included patients who had their appointments cancelled in March 2020 due to the 1st lockdown, and high-risk patients. This particular patient group may have an increased urgency to catch up on screening. This could have contributed to the increased uptake rate of screening in Scotland in the study period. Increase in uptake rates could also be attributed to increased accessibility for patients due to the “work-from-home” model and increased health consciousness due to COVID-19. Neither raw data nor sample size was defined in study and will require future analysis.

Due to the inherent weaknesses of a rapid review, certain limitations are present within the study as explored below. However, this study can be expanded upon by various means (also explored below) to further elucidate the global impact of COVID-19 on breast cancer detection and subsequent care. Other limitations include the COVID-19 pandemic context as an evolving field with fast publication turnovers; more papers could have been published since the review started. This issue could be partially addressed by completing a repeat search with employment of forwards and backwards citation tracking, while including more grey literature sources apart from governmental databases (e.g., private screening databases).

Other limitations included studies had insufficient data for combined analysis regarding COVID-19 waves past December 2020. Additionally, data obtained was cross-sectional instead of cohort-based; we were unable to analyze trends and recovery in breast cancer screening rates and incidence rates over time. Exclusion of non-English language literature was a weakness. Many countries with extensive population-based breast screening programs that were affected by COVID-19 in Europe and Asia were unaccounted for; the inclusion of additional data would be useful to clarify the impact of the pandemic on breast cancer screening program uptake.

In summary, screening volume and uptake rates were generally reduced but many studies showed gains over time even if overall a decline in screening volume observed. These declines were likely due to the first COVID-19 wave where many health care facilities paused non-essential services. Volume and uptake reductions of smaller magnitudes were observed and our data suggest some difference depending on region and health care coverage. Access to screening services may increase marginalization of some vulnerable groups in the USA due to the pandemic and recovery efforts to reduce disparities in access to screening and early diagnosis should be monitored to determine if prevention services need strengthening. Participation and volume are not conclusive endpoints themselves and future work on from registries and other data sources are needed to determine if there has been any impact on incidence, stage and mortality outcomes.

## Data Availability

All data analysed is provided in the manuscript or supplements

## References

Amornsiripanitch, N., Chikarmane, S. A., Bay, C. P. & Giess, C. S. 2021. Patients characteristics related to screening mammography cancellation and rescheduling rates during the COVID-19 pandemic. Clinical Imaging 80, 205–210. DOI: 10.1016/j.clinimag.2021.07.009

Becker, N. V., Moniz, M. H., Tipirneni, R., Dalton, V. K. & Ayanian, J. Z. 2021. Utilization of Women’s Preventive Health Services During the COVID-19 Pandemic. JAMA Health Forum, 2, e211408–e211408. DOI: 10.1001/jamahealthforum.2021.1408

Bessa, J. D. F. 2021. Breast imaging hindered during covid-19 pandemic, in Brazil. Revista de saude publica 55, 8–8. DOI: 10.11606/s1518-8787.2021055003375

Berry, D. A., Cronin, K. A., Plevritis, S. K., Fryback, D. G., Clarke, L., Zelen, M., Mandelblatt, J. S., Yakovlev, A. Y., Habbema, J. D., Feuer, E. J., Cancer, I. & Surveillance Modeling Network, C. 2005. Effect of screening and adjuvant therapy on mortality from breast cancer. N Engl J Med 353, 1784–92. DOI: 10.1056/NEJMoa050518

Campbell, C., Sommerfield, T., Clark, G. R. C., Porteous, L., Milne, A. M., Millar, R., Syme, T. & Thomson, C. S. 2021. COVID-19 and cancer screening in Scotland: A national and coordinated approach to minimising harm. Preventive Medicine 151, 106606. DOI: 10.1016/j.ypmed.2021.106606

Caplan, L. S., May, D. S. & Richardson, L. C. 2000. Time to diagnosis and treatment of breast cancer: results from the National Breast and Cervical Cancer Early Detection Program, 1991-1995. Am J Public Health 90, 130–4. DOI: 10.2105/ajph.90.1.130

Chen, R. C., Haynes, K., Du, S., Barron, J. & Katz, A. J. 2021. Association of Cancer Screening Deficit in the United States With the COVID-19 Pandemic. JAMA Oncology 7, 878–884. DOI: 10.1001/jamaoncol.2021.0884

Chiarelli, A. M., Walker, M. J., Espino-Hernandez, G., Gray, N., Salleh, A., Adhihetty, C., Gao, J., Fienberg, S., Rey, M. A. & Rabeneck, L. 2021. Adherence to guidance for prioritizing higher risk groups for breast cancer screening during the COVID-19 pandemic in the Ontario Breast Screening Program: a descriptive study. CMAJ Open 9, E1205–E1212.

Adherence to guidance for prioritizing higher risk groups for breast cancer screening during the COVID-19 pandemic in the Ontario Breast Screening Program: a descriptive study

DeGroff, A., Miller, J., Sharma, K., Sun, J., Helsel, W., Kammerer, W., Rockwell, T., Sheu, A., Melillo, S., Uhd, J., Kenney, K., Wong, F., Saraiya, M. & Richardson, L. C. 2021. COVID-19 impact on screening test volume through the National Breast and Cervical Cancer early detection program, January–June 2020, in the United States. Preventive Medicine 151, 106559. DOI: 10.1016/j.ypmed.2021.106559

Dennis, L. K., Hsu, C.-H. & Arrington, A. K. 2021. Reduction in Standard Cancer Screening in 2020 throughout the U.S. Cancers 13, 5918. DOI: 10.3390/cancers13235918

Dhahri, A. & Tegene, T. 2021. The impact of COVID-19 on breast cancer care: Delays in screening. Journal of Clinical Oncology 39, 130.

Doubova, S. V., Leslie, H. H., Kruk, M. E., Pérez-Cuevas, R. & Arsenault, C. 2021. Disruption in essential health services in Mexico during COVID-19: an interrupted time series analysis of health information system data. BMJ Global Health 6, e006204. DOI: 10.1136/bmjgh-2021-006204

Eijkelboom, A. H., De Munck, L., Lobbes, M. B. I., Van Gils, C. H., Wesseling, J., Westenend, P. J., Guerrero Paez, C., Pijnappel, R. M., Verkooijen, H. M., Broeders, M. J. M. & Siesling, S. 2021. Impact of the suspension and restart of the Dutch breast cancer screening program on breast cancer incidence and stage during the COVID-19 pandemic. Preventive Medicine 151, 106602. DOI: 10.1016/j.ypmed.2021.106602

Fedewa, S. A., Cotter, M. M., Wehling, K. A., Wysocki, K., Killewald, R. & Makaroff, L. 2021. Changes in breast cancer screening rates among 32 community health centers during the COVID-19 pandemic. Cancer 127, 4512–4515. DOI: 10.1002/cncr.33859

Figueroa, J. D., Gray, E., Pashayan, N., Deandrea, S., Karch, A., Vale, D. B., Elder, K., Procopio, P., Van Ravesteyn, N. T., Mutabi, M., Canfell, K. & Nickson, C. 2021. The impact of the Covid-19 pandemic on breast cancer early detection and screening. Prev Med 151, 106585. DOI: 10.1016/j.ypmed.2021.106585

IARC, 2016. Breast cancer screening: IARC Handbooks of Cancer Prevention, 15^th^ Ed, International Agency for Research on Cancer.

Jidkova, S., Hoeck, S., Kellen, E., Le Cessie, S. & Goossens, M. C. 2022. Flemish population-based cancer screening programs: impact of COVID-19 related shutdown on short-term key performance indicators. BMC Cancer 22, 183. DOI: 10.1186/s12885-022-09292-y

Knoll, K., Reiser, E., Leitner, K., Kögl, J., Ebner, C., Marth, C. & Tsibulak, I. 2022. The impact of COVID-19 pandemic on the rate of newly diagnosed gynecological and breast cancers: a tertiary center perspective. Archives of gynecology and obstetrics 305, 945–953. DOI: 10.1007/s00404-021-06259-5

Lehman, C. D., Mercaldo, S. F., Wang, G. X., Dontchos, B. N., Specht, M. C. & Lamb, L. R. 2021. Screening Mammography Recovery After COVID-19 Pandemic Facility Closures: Associations of Facility Access and Racial and Ethnic Screening Disparities. American Journal of Roentgenology, 1–9.

London, J. W., Fazio-Eynullayeva, E., Palchuk, M. B. & Mcnair, C. 2022. Evolving Effect of the COVID-19 Pandemic on Cancer-Related Encounters. JCO Clinical Cancer Informatics, e2100200. DOI: 10.2214/AJR.21.26890

Losurdo, P., Samardzic, N., Di Lenarda, F., De Manzini, N., Giudici, F. & Bortul, M. 2022. The real-word impact of breast and colorectal cancer surgery during the SARS-CoV-2 pandemic. Updates in Surgery 74(3), 1063–1072. DOI: 10.1007/s13304-021-01212-2

Miller, M. M., Meneveau, M. O., Rochman, C. M., Schroen, A. T., Lattimore, C. M., Gaspard, P. A., Cubbage, R. S. & Showalter, S. L. 2021. Impact of the COVID-19 pandemic on breast cancer screening volumes and patient screening behaviors. Breast Cancer Research and Treatment 189, 237–246. DOI: 10.1007/s10549-021-06252-1

Nyante, S. J., Benefield, T. S., Kuzmiak, C. M., Earnhardt, K., Pritchard, M. & Henderson, L. M. 2021. Population-level impact of coronavirus disease 2019 on breast cancer screening and diagnostic procedures. Cancer 127, 2111–2121. DOI: 10.1002/cncr.33460

OECD (2021), Primary Health Care in Brazil, OECD Reviews of Health Systems, OECD Publishing, Paris, https://doi.org/10.1787/120e170e-en.

Otto, P. M. & Blecher, C. B. 2014. Controversies surrounding screening mammography. Mo Med 111, 439–43.

Page, M. J., Mckenzie, J. E., Bossuyt, P. M., Boutron, I., Hoffmann, T. C., Mulrow, C. D., Shamseer, L., Tetzlaff, J. M., Akl, E. A., Brennan, S. E., Chou, R., Glanville, J., Grimshaw, J. M., Hróbjartsson, A., Lalu, M. M., Li, T., Loder, E. W., Mayo-Wilson, E., Mcdonald, S., Mcguinness, L. A., Stewart, L. A., Thomas, J., Tricco, A. C., Welch, V. A., Whiting, P. & Moher, D. 2021. The PRISMA 2020 statement: an updated guideline for reporting systematic reviews. BMJ 372, 71. DOI: 10.1136/bmj.n71

Peng, S. M., Yang, K. C., Chan, W. P., Wang, Y. W., Lin, L. J., Yen, A. M., Smith, R. A. & Chen, T. H. 2020. Impact of the COVID-19 pandemic on a population-based breast cancer screening program. Cancer 126, 5202–5205. DOI: 10.1002/cncr.33180

Ribeiro, C. M., Correa, F. M. & Migowski, A. 2022. Short-term effects of the COVID-19 pandemic on cancer screening, diagnosis and treatment procedures in Brazil: a descriptive study, 2019-2020. Epidemiol Serv Saude 31, e2021405. DOI: 10.1590/S1679-49742022000100010

Schardt, C., Adams, M. B., Owens, T., Keitz, S. & Fontelo, P. 2007. Utilization of the PICO framework to improve searching PubMed for clinical questions. BMC Med Inform Decis Mak 7, 16. DOI: 10.1186/1472-6947-7-16

Shen, C.-T., Hsieh, H.-M., Chang, Y.-L., Tsai, H.-Y. & Chen, F.-M. 2022. Different impacts of cancer types on cancer screening during COVID-19 pandemic in Taiwan. Journal of the Formosan Medical Association 121(10):1993-2000. DOI: 10.1016/j.jfma.2022.02.006

Sprague, B. L., Lowry, K. P., Miglioretti, D. L., Alsheik, N., Bowles, E. J. A., Tosteson, A. N. A., Rauscher, G., Herschorn, S. D., Lee, J. M., Trentham-Dietz, A., Weaver, D. L., Stout, N. K. & Kerlikowske, K. 2021. Changes in Mammography Use by Women’s Characteristics During the First 5 Months of the COVID-19 Pandemic. J Natl Cancer Inst 113, 1161–1167. DOI: 10.1093/jnci/djab045

Toss, A., Isca, C., Venturelli, M., Nasso, C., Ficarra, G., Bellelli, V., Armocida, C., Barbieri, E., Cortesi, L., Moscetti, L., Piacentini, F., Omarini, C., Andreotti, A., Gambini, A., Battista, R., Dominici, M. & Tazzioli, G. 2021. Two-month stop in mammographic screening significantly impacts on breast cancer stage at diagnosis and upfront treatment in the COVID era. ESMO Open 6, 100055. DOI: 10.1016/j.esmoop.2021.100055

Tricco, A. C., Antony, J., Zarin, W., Strifler, L., Ghassemi, M., Ivory, J., Perrier, L., Hutton, B., Moher, D. & Straus, S. E. 2015. A scoping review of rapid review methods. BMC Med 13, 224. DOI: 10.1186/s12916-015-0465-6

Unger-Saldaña, K., Guadiamos, MC., Vega, AMD., Anderson, B., Romanoff, R,. 2020. Delays to diagnosis and barriers to care for breast cancer in Mexico and Peru: A Cross Sectional Study. The Lancet Global Health, 8(Special Issue). Available at: https://doi.org/10.1016/s2214-109x(20)30157-1.

Walker, M. J., Meggetto, O., Gao, J., Espino-Hernández, G., Jembere, N., Bravo, C. A., Rey, M., Aslam, U., Sheppard, A. J., Lofters, A. K., Tammemägi, M. C., Tinmouth, J., Kupets, R., Chiarelli, A. M. & Rabeneck, L. 2021. Measuring the impact of the COVID-19 pandemic on organized cancer screening and diagnostic follow-up care in Ontario, Canada: A provincial, population-based study. Preventive Medicine 151, 106586. DOI: 10.1016/j.ypmed.2021.106586

Zhao, J., Mao, Z., Fedewa, S. A., Nogueira, L., Yabroff, K. R., Jemal, A. & Han, X. 2020. The Affordable Care Act and access to care across the cancer control continuum: A review at 10 years. CA Cancer J Clin 70, 165–181. DOI: 10.3322/caac.21604

## Web References

BreastScreen Aoteroa. 2021. Breast Screening [Online]. Available: https://www.timetoscreen.nz/breast-screening/ [Accessed 20/04/2022].

BreastScreen Australia. 2021. Breast Screening [Online]. Available: https://www.aihw.gov.au/reports/cancer-screening/cancer-screening-and-covid-19-in-australia/data [Accessed 20/04/2022].

IARC, 2022. Cancer Today [Online]. IARC. Available: https://gco.iarc.fr/today/online-analysis-map?v=2020&mode=population&mode_population=continents&population=900&populatio ns=900&key=crude_rate&sex=2&cancer=20&type=2&statistic=1&prevalence=1&populatio n_group=0&ages_group%5B%5D=0&ages_group%5B%5D=17&nb_items=10&group_cancer=1&include_nmsc=0&include_nmsc_other=0&projection=natural-earth&color_palette=default&map_scale=quantile&map_nb_colors=5&continent=0&show_r anking=0&rotate=%255B10%252C0%255D [Accessed 11/04/2022].

Critical appraisal tools (2022) Joanna Briggs Institute. Available at: https://jbi.global/critical-appraisal-tools (Accessed: December 8, 2022).

McKenzie, J. 2022. Grey literature: What it is & how to find it [Online]. Available: https://www.lib.sfu.ca/help/research-assistance/format-type/grey-literature#:~:text=Grey%20literature%20is%20information%20produced,urban%20plans%2C%20and%20so%20on. [Accessed 20/04/2022].

Medicaid.gov. 2022. Medicaid Eligibility [Online]. Available: https://www.medicaid.gov/ [Accessed 20/04/2022].

NHS England. 2021. Breast Screening Programme: National statistics, Official statistics. NHS Digitial: NHS England. Available: https://digital.nhs.uk/data-and-information/publications/statistical/breast-screening-programme [Accessed 20/04/2022].

OECD. 2021. Breast cancer screening (mammography), survey data and programme data [Online]. In: OECD, OECD Health Statistics 2021. Available: https://stats.oecd.org/FileView2.aspx?IDFile=eb5acd7d-2445-401a-b624-62fcdad85091 [Accessed 20/04/2022]

PAHO, 2020. Mexico Cancer Country Profile. In: Organisation, PAHO, Cancer Country Profile 2020 [Online], World Health Organisation. Available: https://www3.paho.org/hq/index.php?option=com_docman&view=download&category_slug=4-cancer-country-profiles-2020&alias=51536-mexico-cancer-profile-2020&Itemid=270&lang=en [Accessed 20/04/2022].

WHO. 2022. Cancer Fact Sheet [Online]. World Health Organisation. Available: https://www.who.int/news-room/fact-sheets/detail/cancer [Accessed 20/04/2022].

Worldometer. 2022. COVID-19 Coronavirus Pandemic Meter [Online]. Available: https://www.worldometers.info/coronavirus/ [Accessed 20/04/2022].

WHO, 2022. Breast cancer. [online] Who.int. Available at: <https://www.who.int/news-room/fact-sheets/detail/breast-cancer> [Accessed 20/04/2022].

Yucatan Times. 2021. More and more Yucatecan women are visiting IMSS for breast cancer prevention exams. Yucatan Times. [online] Available at: <https://www.theyucatantimes.com/2021/10/more-and-more-yucatecan-women-are-visiting-imss-for-breast-cancer-prevention-exams/> [Accessed 20/04/2022]

